# A Particle-Based COVID-19 Simulator with Contact Tracing and Testing

**DOI:** 10.1101/2020.12.07.20245043

**Authors:** Askat Kuzdeuov, Aknur Karabay, Daulet Baimukashev, Bauyrzhan Ibragimov, Huseyin Atakan Varol

## Abstract

**Goal:** The COVID-19 pandemic has emerged as the most severe public health crisis in over a century. As of January 2021, there are more than 100 million cases and 2.1 million deaths. For informed decision making, reliable statistical data and capable simulation tools are needed. Our goal is to develop an epidemic simulator that can model the effects of random population testing and contact tracing.

**Methods:** Our simulator models individuals as particles with the position, velocity, and epidemic status states on a 2D map and runs an SEIR epidemic model with contact tracing and testing modules. The simulator is available on GitHub under the MIT license.

**Results:** The results show that the synergistic use of contact tracing and massive testing is effective in suppressing the epidemic (the number of deaths was reduced by 72%).

**Conclusions:** The Particle-based COVID-19 simulator enables the modeling of intervention measures, random testing, and contact tracing, for epidemic mitigation and suppression.

**Impact Statement:** Our particle-based epidemic simulator, calibrated with COVID-19 data, models each individual as a unique particle with a location, velocity, and epidemic state, enabling the consideration of contact tracing and testing measures.

## I. INTRODUCTION

**C**OVID-19 has emerged as the most threatening health care crisis in over a century, spreading rapidly throughout the world. The first cases were identified in Wuhan, China (December 2019), and within months the World Health Organization declared the disease to be a pandemic (March 11, 2020) [1]. The propagation of the virus has increased rapidly in spite of unprecedented government interventions intended to suppress and mitigate the spread; as of January 26, 2021, more than 100.3 million cases with 2.15 million deaths have been reported [2]. However, the actual number of cases is likely, significantly higher because of limited testing and the high percentage of asymptomatic cases [3].

The propagation was accelerated by multiple convergent factors, led first by the fact that it is a novel coronavirus to which neither is there existing resistance amongst the population, nor an effective vaccine against. Different vaccines against COVID-19 are being developed at a historic rate, but their effectiveness and safety are still an open question [4]. Until a vaccine that can be distributed on a massive scale emerges, the only recourse to retard the spread of the disease is what is known as non-pharmaceutical interventions (NPIs), such as quarantines, travel restrictions [5], online education [6], and large-scale virus testing with comprehensive contact tracing of infected individuals [7],[8].

The lockdown policies slow down the propagation of the disease, but they also inflict substantial social and economic damage [9],[10], and, when done indiscriminately, the negative effects are non-trivial; interdiction measures should be enacted in ways that mitigate disease spread while minimizing negative effects. One way to achieve a more nuanced approach, with less collateral damage, is the use of computer models and simulations.

One of the earliest epidemic models, Susceptible-Infected-Recovered (SIR) [11], divides the population into three compartments. In the first compartment, susceptible (S) individuals are vulnerable but not infected. In the second compartment, infected (I) individuals are infected and capable of transmitting the disease to the susceptible individuals. The last compartment, recovered (R) contains individuals who have overcome the disease. The recovered individuals are assumed to have acquired some level of immunity to the disease, thus they have a lower probability of reinfection, compared to the susceptible individuals.

The compartmental models have been widely used in modeling the spread of COVID-19 [12]–[14]. Despite their popularity, the compartmental models have several limitations because of the simplifying assumptions that do not represent the actual viral propagation. For instance, the compartmental models do not consider each individual separately. Therefore, the mobility and current epidemic state of each individual, the moments of getting infected and recovered are omitted. As a result, the contact tracing and testing policies at the individual level cannot be implemented.

A number of works on epidemic simulation at the individual level can be found in the literature. For instance, a particle-modeling approach based on the Monte Carlo algorithm was developed to simulate the spread of COVID-19 [15]. The results show that periodic lockdown, and strict social distancing might help to keep the infection rate under control. A stochastic agent-based model was employed for simulating COVID-19 in France [16]. An SEIR agent-based model was implemented to analyze different social distancing interventions in [17]. An agent-based simulation was implemented by Bicher et al. [18] to estimate the effectiveness of contact tracing policies.

In this work, we have developed a particle-based simulator which models each individual as a unique particle with a location, velocity, and epidemic state. We provide a demo video in the supplementary materials that illustrates the particles’ motion, transitions between epidemic states, and effects of the contact tracing and testing modules. To the best of our knowledge, it is the first particle-based SEIR model with contact tracing and testing, that was calibrated with actual COVID-19 data. The particles move randomly on a square 2D map, and become infected if they enter the proximity of an infectious particle closer than a predefined physical distance. The contact tracing module is based on the use of a mobile app and stores the list of contacts for each particle such that if a particle is determined to be infected, then all particles on the list are quarantined or isolated. The testing module simulates the massive random testing of the population. The module considers test sensitivity and specificity [19]. This way, the simulator is able to yield different scenarios and mitigation policies.

The rest of the paper is organized as follows: in Section II, we introduce our method of implementing the particle simulator. We calibrate the particle simulator using real COVID-19 data in Section III. Afterward, we simulate scenarios, using different contact tracing ratios and the daily number of tests per thousand people. In Section IV, we discuss the simulation results, and the Section V concludes our work.

## II. MATERIALS AND METHODS

### A. Particle Model

In this work, each individual is considered as a particle *p* and modeled as

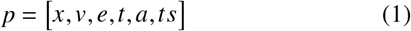

where *x* ∈ ℝ^2^ is the position of a particle on the map; *v* ∈ ℝ^2^ is the particle velocity; *e* is the epidemic state of the particle (i.e., susceptible, exposed, infected, recovered, dead, quarantined, isolated, or severely infected); *t* is the time of the particle in the current epidemic state, and it is incremented by the sampling time Δ*t* at each iteration of the simulation; *a* denotes the availability of the contact tracing application; *ts* is the COVID-19 test result of the particle.

The current position and velocity of *n* particles are stored in matrices ***X*** ∈ ℝ^*nx*2^ and ***V*** ∈ ℝ^*nx*2^, and constrained by −1 ≤ *x*_*i j*_ ≤ 1,− *v*_*max*_ ≤ *v*_*ij*_ ≤ *v*_*max*_ for all particles *i* = 1, …, *n* and two dimensions *j* = 1, 2. The initial values are set randomly by taking into account the imposed constraints. The velocity matrix *V* ∈ ℝ^*nx*2^ is updated at each iteration *κ* (1≤ *κ* ≤*T*/Δ*t*) in the simulation as

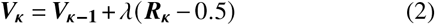

where ***R***_***K***_ ∈ ℝ^*nx*2^ is a matrix of uniformly distributed random numbers in the interval [0, 1], and *λ* is a momentum that allows to control velocity change. Velocities are reset to zero if they exceed the maximum allowed speed *v*_*max*_. In addition, dead, quarantined, isolated, and severely infected (hospitalized) particles are considered to be stationary (i.e., their velocities are set to zero). The ***R***_***κ***_ is normalized to the [-0.5, 0.5] range. Otherwise, the velocity keeps increasing until the maximum velocity *v*_*max*_ is reached. Also, it is important to note that the maximum allowed speed of particles *v*_*max*_ impacts the rate of the epidemic spread among the population. In order to explore this relation, we have provided simulation results for different values of the maximum velocity in the supplementary materials.

Using (2), the position matrix ***X*** ∈ ℝ^*nx*2^ is updated as

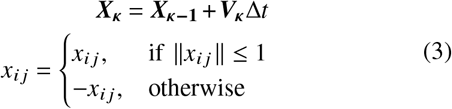

such that if particles reach one of the borders of the map, they appear on the opposite side. This is necessary to keep all the particles inside the map.

### B. Particle-Based SEIR Simulator

The particle-based simulator consists of four superstates: Susceptible (**S**^*s*^), Exposed (**E**^*s*^), Infected (**I**^*s*^), and Recovered (**R**^*s*^). Transitions between states are shown in Fig. 1. The Exposed superstate (**E**^*s*^) is composed of Exposed (**E**) and Quarantined (**Q**) states. The Quarantined (**Q**) state consists of True Quarantined (**TQ**) and False Quarantined (**FQ**) substates. Similarly, the Infected superstate (**I**^*s*^) consists of Infected (**I**), Isolated (**Iso**), and Severely Infected (**SI**) states. The Isolated (**Iso**) state has two substates: True Isolated (**TIso**) and False Isolated (**FIso**). To avoid confusions between the superstates (e.g., **E**^*s*^) and states (e.g., **E**), we will further refer to the states only. Also, when particles transition from the current superstate to another, their time *t* in the current superstate is reset to zero and starts over in the new superstate. The time is not restarted if transitions occur between states of the same superstate.

**Fig. 1.**
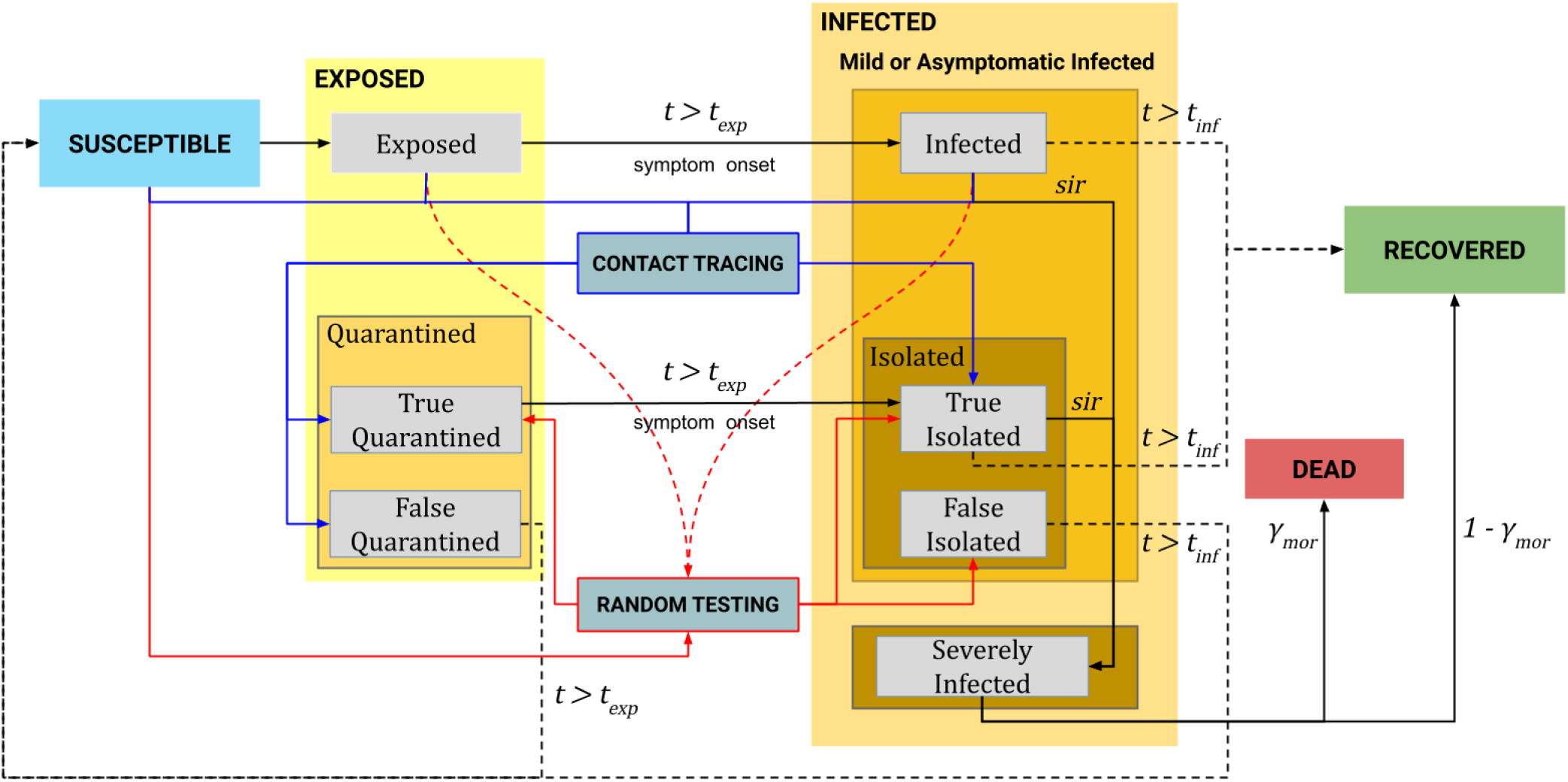
The statechart of the particle-based SEIR epidemic simulator.

At the beginning of the simulation, we randomly assign a small number of *n*_*e*_ particles to the Exposed state, while other *n*− *n*_*e*_ particles are in the Susceptible state. The list and description of simulation parameters are given in Table I. At each iteration, susceptible particles become exposed when they come into contact with contagious particles (**I, E, TQ, TIso**, and **SI**). The contact occurs when the distance between the particles becomes less than the contact threshold x_*thr*_. The disease transmission probability for an infected particle is equal to one while for other contagious particles are defined by the parameters ϵ_*exp*_for **E**, ϵ_*qua*_ for **TQ** and **TIso**, and ϵ_*sev*_for **SI** (in the range from 0 to 1). The exposed particles transition to the Infected state after *t*_*exp*_ days. Then, some portion of the infected particles become severely infected according to the rate of daily transition to Severely Infected, *sir*, while the others move to the Recovered state after *t*_*inf*_ days.

**TABLE I.**
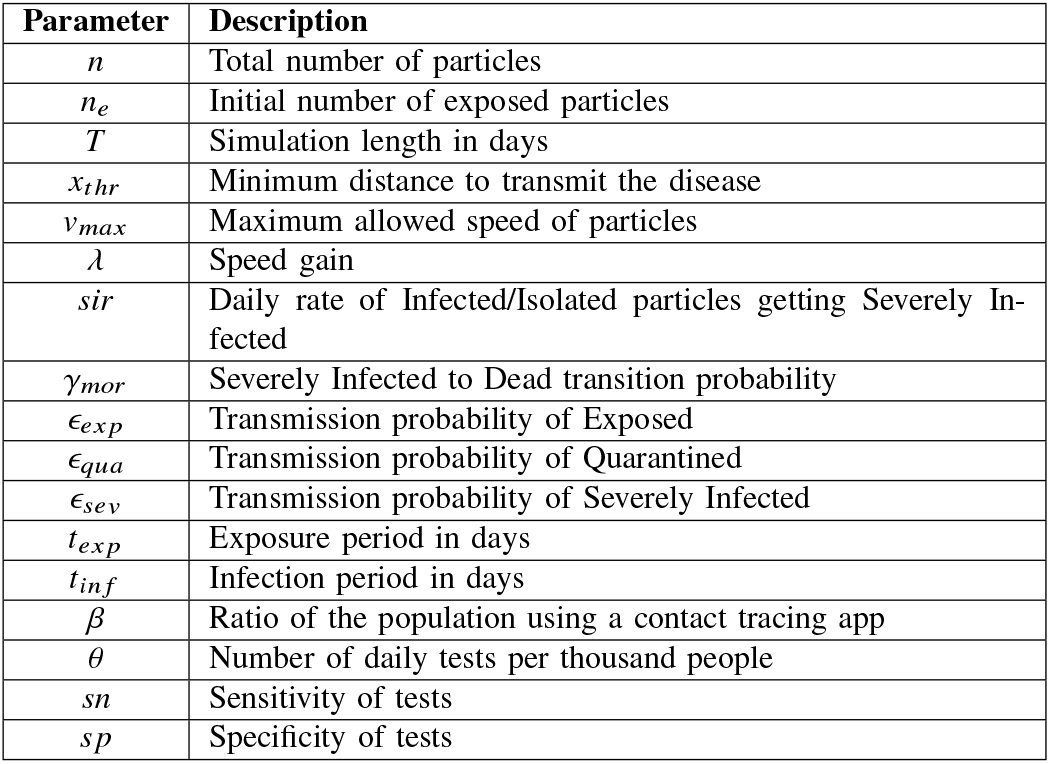
List of simulation parameters and their descriptions.

The testing module randomly tests particles in the Susceptible, Exposed, and Infected states at each iteration of the simulation and changes their test status *ts* accordingly. The number of daily tests per thousand people is set by the parameter *θ*. The test sensitivity and specificity are defined by parameters *sn* and *sp*, respectively. The exposed and infected particles that were correctly detected by the test are sent to the True Quarantined and True Isolated states, respectively. The susceptible particles that were tested false positive are sent to the False Isolated state. The infected particles in the True Isolated state transition to the Severely Infected state according to the *sir* rate, while the other particles in this state recover after *t*_*inf*_ days. The False Isolated particles do not transition to the Severely Infected state (since they are actually not infected) and, therefore, transition back to the Susceptible state after *t*_*inf*_ days. Particles in the Severely Infected state die according to the mortality rate *γ*_*mor*_. The rest transfer to the Recovered state.

The proportion of the population using a contact tracing app is defined by the parameter *β* With the help of the app, the contact tracing module stores a list of contacted particles for each particle and the corresponding contact instant. If a certain particle is determined to be infected (i.e., the test result is positive), then its list of contacted particles in the last *t*_*inf*_ days is extracted and tested. If a contact of the true positive tested particle is in the Exposed state, then the contact is sent to the True Quarantined state. If the contact is in the Infected state, then it is sent to the True Isolated state. The contacts of the false positive tested particle can only be susceptible particles. Therefore, they are sent to the False Quarantined state. Then, particles in the True Quarantined state move to the True Isolated state after *t*_*exp*_ days. Particles in the False Quarantined state go back to the Susceptible state. A detailed explanation, in the form of equations, for the epidemic state transitions can be found in the supplementary materials.

## III. RESULTS

### A. Particle-Based SEIR Simulation of Lecco

In this section, we simulate the epidemic in the province of Lecco, Italy. Lecco is located in the Lombardy region, which was the epicenter of the COVID-19 outbreak in Italy. We chose this province, because the epidemic timeline of Lombardy is well-established, and official statistics of daily epidemic data for the region and its provinces have been shared with public since February 24, 2020 [20]. The timeline of important events and NPIs listed in Table II was used to calibrate the model parameters and imitate the real pattern of epidemic spread.

**TABLE II.**
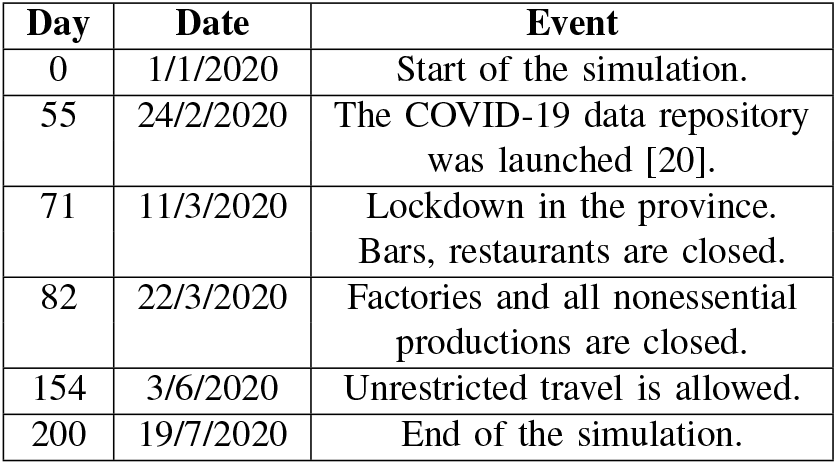
Major events and NPIs in Lecco province during the COVID-19 epidemic.

| Day | Date | Event |
| --- | --- | --- |
| 0 | 1/1/2020 | Start of the simulation. |
| 55 | 24/2/2020 | The COVID-19 data repository was launched [20]. |
| 71 | 11/3/2020 | Lockdown in the province. Bars, restaurants are closed. |
| 82 | 22/3/2020 | Factories and all nonessential productions are closed. |
| 154 | 3/6/2020 | Unrestricted travel is allowed. |
| 200 | 19/7/2020 | End of the simulation. |

The total number of particles *n* was set to 337,000 (the population of Lecco). The other parameters of the simulation are provided in Table III. According to the seroprevalence survey results presented on August 3, 2020 by the Italian Ministry of Health [21], it was estimated that 1,482,000 people had encountered the virus in Italy, which is six times greater than the officially registered cases. Therefore, we assumed that the actual number of total cases in Lecco are also six times greater than the registered cases. Because the daily deaths for the provinces are not available in [20], we used a proportional amount from the total deaths officially reported for the Lombardy region.

We started the simulation on January 1, 2020, based on the results in [22], with initially 10 exposed particles. The length of the simulation was set to 200 days. Regarding the parameters of the testing module, there were no official information on the used test kits and on the number of daily tests per thousand people for Lecco. Therefore, we estimated the daily tests per thousand people *θ* at 0.5 for the considered period in the simulation, based on the testing data for the whole Italy [23]. For the test sensitivity *sn* and specificity *sp*, we used the values of most commonly manufactured tests kits [24]. In order to imitate the lock-down in Lecco, we decreased the maximum speed of particles *v*_*max*_ and *λ* according to the timeline of events, and when the lock-down was lifted on June 3, 2020, we returned them to their initial values, assuming that people started traveling as usual. However, the contact threshold *x*_*thr*_ was decreased slightly considering that the population started wearing masks and keeping physical distancing.

**TABLE III.** Simulation Parameters for Lecco.

| $v_{max}$ (Days) | $\lambda$ (Days) | $x_{thr}$ (Days) | $\epsilon_{exp}$ | $\epsilon_{qua}$ | $\epsilon_{sev}$ | $t_{exp}$ |
| --- | --- | --- | --- | --- | --- | --- |
| 0.02 (0-55) | 0.002 (0-55) |  |  |  |  |  |
| 0.012 [55-71] | 0.0012 [55-71] | 8.6e-5 (0-154) |  |  |  |  |
| 0.006 [71-82] | 0.0006 [71-82] | 6.9e-5 [154-200] | 0.7 | 0.3 | 0.3 | 5 |
| 0.004 [82-154] | 0.0004 [82-154] |  |  |  |  |  |
| 0.02 [154-200] | 0.002 [154-200] |  |  |  |  |  |
| $t_{inf}$ | $sir$ | $\gamma_{mor}$ | $\beta$ | $\theta$ | $sn$ | $sp$ |
| 14 | 0.02 | 0.15 | 0 | 0.5 | 0.95 | 0.99 |

The averaged results of ten simulations are shown in Fig. 2 with the standard deviations for the total cases and deaths. According to the reported data for the province of Lecco [20], new daily cases increased significantly starting from the middle of March and remained high until the middle of April. This is also observed in our simulation results. Thus, we conclude that our simulator predicted the peak of the epidemic correctly. Also, in the simulation results, the average number of total cases were approximately five times higher than the reported numbers, which is similar to the results of the seroprevalence test for the whole Italy.

**Fig. 2.**
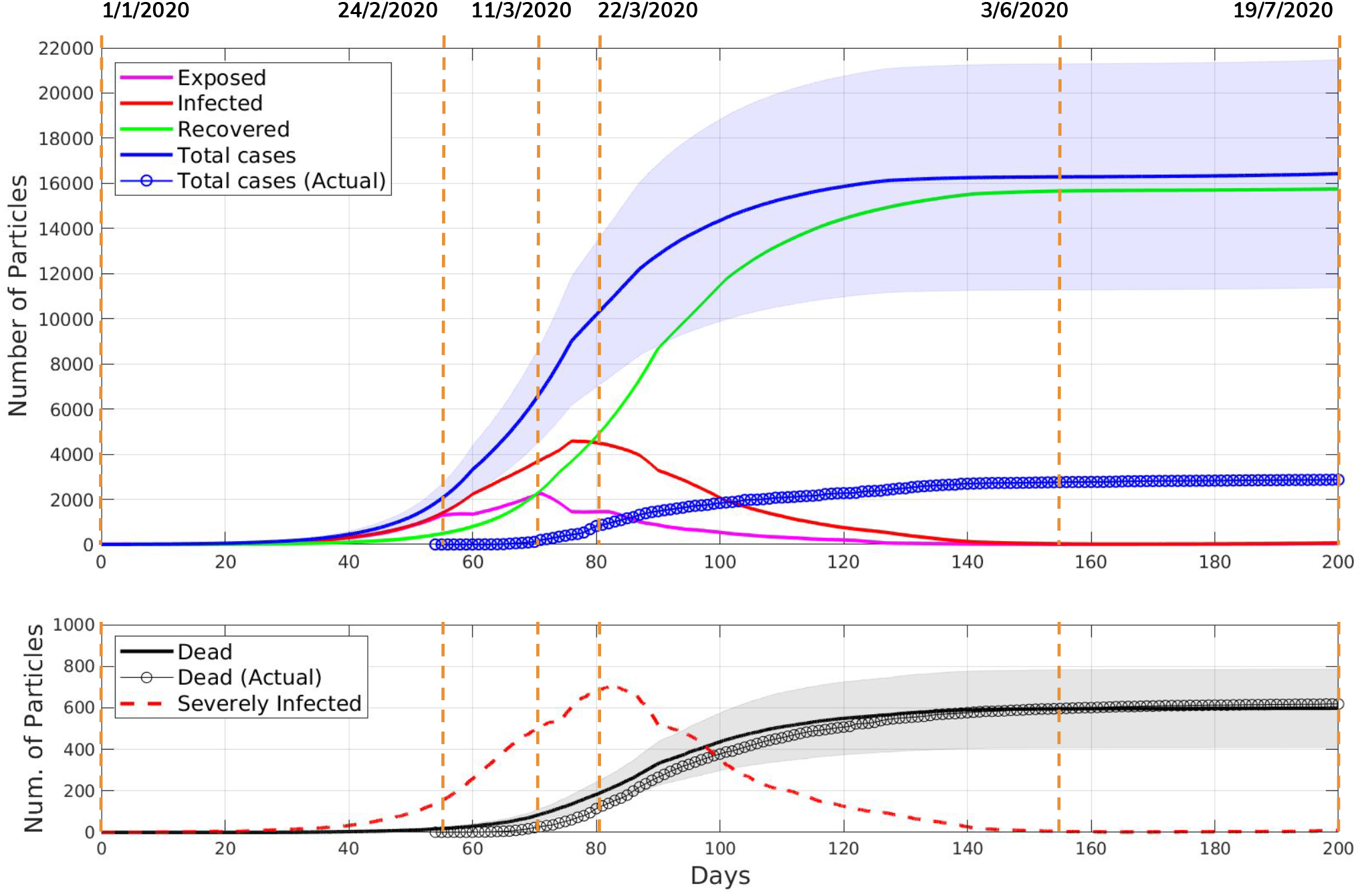
The averaged results of ten simulations for the province of Lecco. The upper plot shows the states of the epidemic simulation versus time. The dates of important NPIs are shown by vertical dashed lines and listed in Table II. One standard deviation around the average Total case curve is shaded in blue. The bottom plot compares the average number of deaths in the simulation with the actual number of deaths due to COVID-19. This plot also shows the simulation results for the number of severely infected particles as well. One standard deviation around the average Dead state curve is shaded in grey.

### B. Simulations with Contact Tracing and Testing Modules

In this section, we analyze the impact of randomly testing the population and tracing contacts of positive tested individuals in reducing the spread of the epidemic. First, we considered the case of massive testing the population without the contact tracing policy. Thus, we set to *β* zero and conducted simulations for different values of *θ* = {0, 5, 10, 15, 20}. The results of the simulations are shown in the top row of Fig. 3. According to the results in Figs. 3a and 3b, the number of particles in the Isolated and Quarantined states increases with the increased value of *θ*. Consequently, the number of infected particles, at the peak of the epidemic, reduced gradually from 4,925 (*θ* = 0) to 3,633 (*θ* = 5), 2,926 (*θ* = 10), 2,428 (*θ* = 15), and to 1,841 (*θ* = 20) (see Fig. 3c).

**Fig. 3.**
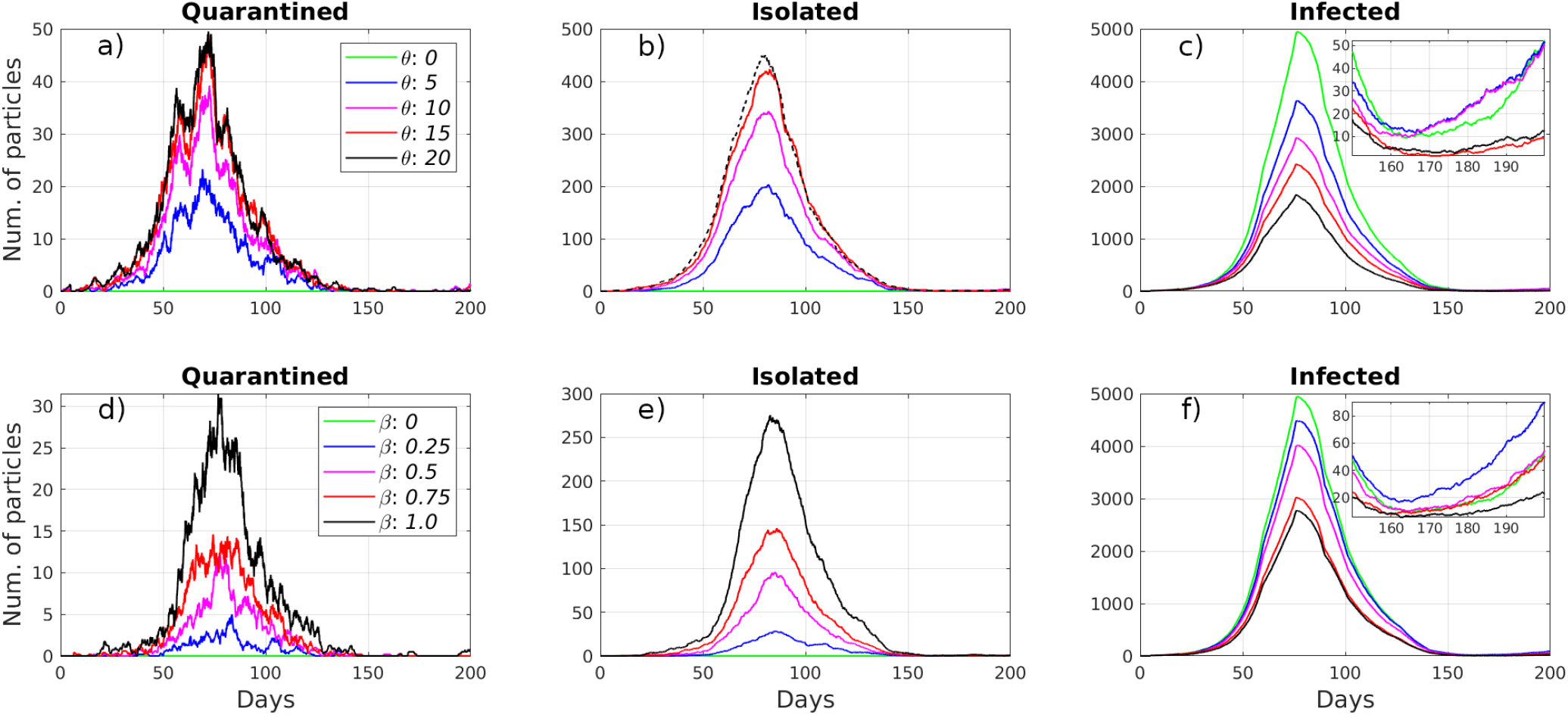
Simulation results of using only the random testing module: a, b, c, and using only the contact tracing module: d, e, f.

Similarly, we examined the effect of the contact tracing policy without the randomized testing of the population. We set *θ* to zero and simulated with different values of *β* = {0.0, 0.25, 0.5, 0.75, 1.0}. In this case, we traced particles that were in contact only with severely infected particles (i.e., hospitalized) because infected and exposed particles can be found by randomly testing the population. The results of the simulations are shown in the bottom row of Fig. 3. According to Figs. 3d and 3e, the number of particles in the Isolated and Quarantined states increased with the increased value of β. As a result, the number of infected particles at the peak of the epidemic, decreased from 4,925 (*β=* 0) to 4,487 (*β*= 0.25), 4,019 (*β*= 0.*5)*, 3,030 (*β*= 0.75), and to 2,273 (*β*= 1.0) (see Fig. 3f).

Next, we considered the utilization of concurrent contact tracing and massive testing. The considered numbers of daily tests per thousand people and the contact tracing ratios were *θ* = {0, 10, 20} and ={0,0.5, 1.0}, respectively. The results in Figs. 4a and 4b for *θ* = 10 show that the enabled contact tracing module increases the number of isolated and quarantined particles. However, for *θ* = 20, the numbers decrease with the increased contact tracing ratios. The reason is that a higher number of tests allow to find infected and exposed particles at the beginning of the epidemic faster, and additional contact tracing accelerates this even further. Therefore, at the peak of the epidemic, we get a lower number of infected and exposed particles, and, as a result, lower number of isolated and quarantined particles. Nevertheless, in both cases, the contact tracing module decreased the number of infected and exposed particles further, as shown in Figs. 4c and 4d. The number of infected particles, at the peak of the epidemic, dropped from 2,926 (*θ* = 10, = 0) to 2,360 (*θ* = 10, = 0.5) and 2,071 (*θ* = 10, = 1.0). The effect became more pronounced with increased daily testing. Specifically, the number of infected particles reduced from 1,841 (*θ* = 20, = 0) to 1,547 (*θ* = 20, β= 0.5) and to 1,004 (*θ* = 20, = 1.0).

**Fig. 4.**
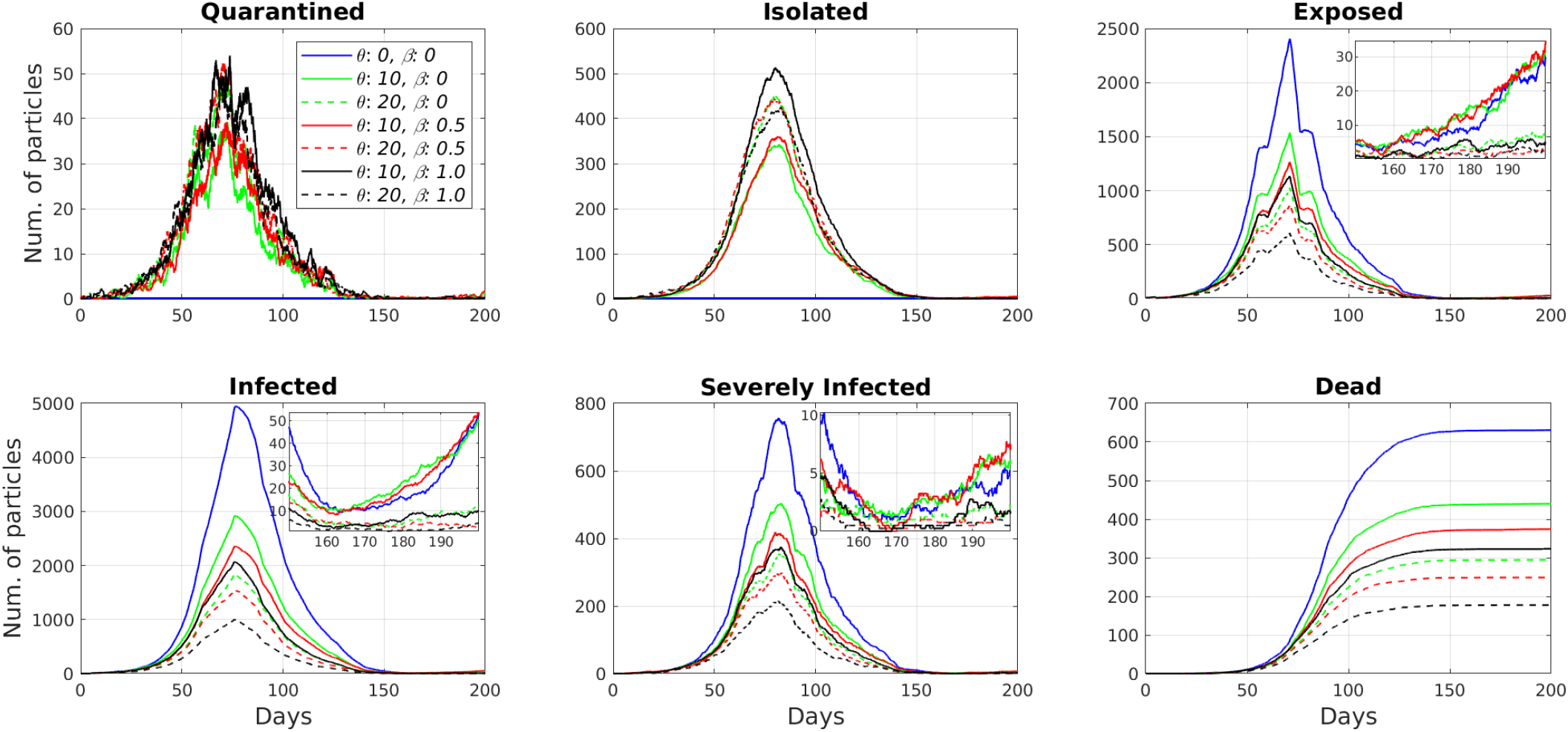
Simulation results for the different combinations of random testing and contact tracing modules.

## IV. DISCUSSION

The simulation results showed that random testing is more efficient compared to the contact tracing module in reducing the number of infected particles when they are used separately. When we use contact tracing without random testing, we trace the contacts of only severely infected particles. The exposed and infected particles continue infecting susceptible particles until they transition to the Severely Infected state. Therefore, the contact tracing module has a limited effect alone. Also, if we observe the zoomed subplots in Figs. 3c and 3f, we see that neither method prevents the second wave of the epidemic after the lifting of restrictions on June 3, 2020.

On the other hand, the synergistic use of two modules showed the most effective results. Namely, the massive testing strategies with (*θ* = 10, = 0) and (*θ* = 20, = 0) reduced the total number of deaths from 630 (*θ* = 0, = 0) to 440 (30%) and 294 (53%), respectively. Then, in the simulations with half of the population using the contact tracing app (*β* = 0.5), massive testing *θ* = 10 and *θ* = 20 reduced the total number of deaths up to 40% (374 deaths) and 60% (249), respectively. The reduction in the number of deaths reached its maximum with ubiquitous contact tracing (β= 1.0). The simulations return 323 deaths (48% reduction) for (*θ* = 10, = 1.0) and 177 deaths (72% reduction) for (*θ* = 20, = 1.0). Also, the zoomed subplots in Fig. 4c, d, and e illustrate that the synergistic use of the two modules prevents the second wave of the epidemic. These results reveal the importance of the immediate isolation of contacts of positive tested particles in preventing the spread of the epidemic.

Even though the modeling of individuals as particles enables the implementation of contact tracing and massive testing, our simulator has several limitations. Firstly, our map is a unit square with the individuals distributed randomly and moving freely without obstacles. In the real world, there are obstacles, for example, buildings and geographic objects such as rivers and mountains. Moreover, the population density differs substantially in different regions of a city or province. The probability of infection is also lower in open spaces than in confined ones. Secondly, the mortality rate for COVID-19 is age and gender dependent [25]. Our particles are identical; demographics properties such as age and gender are not considered. Presumably, the simulator can be enriched by adding demographic profiles and associated risk probabilities for more realistic transitions from the Severely Infected to Dead state. However, this would increase the number of simulation parameters significantly and make the model calibration harder. Thirdly, the simulator does not consider the interaction networks of individuals. Though, in reality, individuals have a number of contacts with whom they interact regularly (e.g., family members, colleagues, and close friends).

## V. CONCLUSION

We developed a particle-based SEIR simulator with contact tracing and testing. The main advantage of our simulator, as compared to the compartmental SEIR model, is that it models each individual as a particle, thus enabling a more realistic simulation of disease propagation, and the impact of intervention strategies for suppression and mitigation. We demonstrated that the simulator can model a real epidemic in accordance with the actual timeline of events and deployment of intervention strategies. We also investigated the impact of contact tracing and testing strategies on the propagation of the disease; the results showed that the most effective approach is an aligned strategy of testing and contact tracing. In future works, the particle-based simulator can be used to simulate the spread of the disease in more confined settings, such as inside buildings (e.g., airports, schools, malls, etc.) by modeling the moving particles according to the specific building layouts.

## Data Availability

As supplementary materials, we provide the source code of the simulator, equations for the transitions between the epidemic states, and a video that illustrates the effectiveness of using random testing and contact tracing strategies.

https://www.youtube.com/watch?v=BJfjmWfi6ac&feature=youtu.be

https://github.com/IS2AI/Particle-Based-COVID19-Simulator

## Supplementary Materials

We implemented the simulator in MATLAB R2020. The source code was uploaded to GitHub^1^ under MIT license.

We also provide a video that illustrates a random motion of a single particle, and also the visualization of particles motion on a 2D map with the corresponding epidemic state transitions for three different scenarios. The first scenario for *θ* = 0, β = 0 shows that the epidemic was suppressed in Lecco only because of the complete lock-down. The second scenario for *θ* = 20, β = 0 illustrates that the lock-down with the additional random testing strategy can reduce the number of infected particles. However, the previous two scenarios also show that neither method ensures the prevention of the second wave of the epidemic. The third scenario with *θ* = 20, β = 1 shows the efficiency of the additional contact tracing strategy. It significantly reduces the number of infected particles and also allows to prevent the second wave of the epidemic.

As an additional example, we have performed a simulation for the canton of Geneva, Switzerland. In this way, we illustrated that the model calibrated for the province of Lecco can be used to predict the epidemic of a different region by adjusting the region-specific parameters such as population, timeline of policies, and COVID-19 statistics.

## Footnotes

1 https://github.com/IS2AI/Particle-Based-COVID19-Simulator

## Notes

### Competing Interest Statement

The authors have declared no competing interest.

### Funding Statement

No external funding was received.

### Author Declarations

This work did not require IRB approval.

## References

[1] WHO. (2020) Timeline of WHO’s response to COVID-19. Last accessed on 2020-10-2: https://www.who.int/news-room/detail/29-06-2020-covidtimeline.

[2] WHO. (2020) Coronavirus disease (COVID-19) outbreak situation. Last accessed on 2020-10-2: “https://www.who.int/emergencies/diseases/novel-coronavirus-2019”.

[3] D. Oran and E. Topol, “Prevalence of asymptomatic SARS-CoV-2 infection,” Annals of Internal Medicine, vol. 173, no. 5, pp. 362–367, 2020, pMID: 32491919. [Online]. Available: https://doi.org/10.7326/M20-3012

[4] S. P. Kaur and V. Gupta, “COVID-19 Vaccine: A comprehensive status report,” Virus Research, vol. 288, pp. 198 114–198 114, Oct 2020. [Online]. Available: https://doi.org/10.1016/j.virusres.2020.198114

[5] M. U. G. Kraemer, C.-H. Yang, B. Gutierrez, C.-H. Wu, B. Klein, D. M. Pigott et al., “The effect of human mobility and control measures on the COVID-19 epidemic in China,” Science, vol. 368, no. 6490, pp. 493–497, 2020. [Online]. Available: https://science.sciencemag.org/content/368/6490/493

[6] R. M. Viner, S. J. Russell, H. Croker, J. Packer, J. Ward, C. Stansfield et al., “School closure and management practices during coronavirus outbreaks including COVID-19: A rapid systematic review,” The Lancet Child & Adolescent Health, vol. 4, no. 5, pp. 397 – 404, 2020. [Online]. Available: http://www.sciencedirect.com/science/article/pii/S235246422030095X

[7] D. Lee and J. Lee, “Testing on the move: South Korea’s rapid response to the COVID-19 pandemic,” Transportation Research Interdisciplinary Perspectives, vol. 5, p. 100111, 2020. [Online]. Available: http://www.sciencedirect.com/science/article/pii/S2590198220300221

[8] V. J. Lee, C. J. Chiew, and W. X. Khong, “Interrupting transmission of COVID-19: Lessons from containment efforts in Singapore,” Journal of Travel Medicine, vol. 27, no. 3, 2020. [Online]. Available: https://doi.org/10.1093/jtm/taaa039

[9] G. Bonaccorsi, F. Pierri, M. Cinelli, A. Flori, A. Galeazzi, F. Porcelli et al., “Economic and social consequences of human mobility restrictions under COVID-19,” Proc. of the National Academy of Sciences, vol. 117, no. 27, pp. 15 530–15 535, 2020. [Online]. Available: https://www.pnas.org/content/117/27/15530

[10] M. Nicola, Z. Alsafi, C. Sohrabi, A. Kerwan, A. Al-Jabir, C. Iosifidis, M. Agha, and R. Agha, “The socio-economic implications of the coronavirus pandemic (COVID-19): A review,” International Journal of Surgery, vol. 78, pp. 185–193, 2020. [Online]. Available: http://www.sciencedirect.com/science/article/pii/S1743919120303162

[11] W. O. Kermack and A. G. McKendrick, “A contribution to the mathematical theory of epidemics,” Proc. of the Royal Society of London. Series A, vol. 115, no. 772, pp. 700–721, 1927. [Online]. Available: http://www.jstor.org/stable/94815

[12] A. Kuzdeuov, D. Baimukashev, A. Karabay, B. Ibragimov, A. Mirza-khmetov, M. Nurpeiissov, M. Lewis, and H. A. Varol, “A network-based stochastic epidemic simulator: Controlling COVID-19 with regionspecific policies,” IEEE Journal of Biomedical and Health Informatics, vol. 24, no. 10, pp. 2743–2754, 2020.

[13] Z. Yang, Z. Zeng, K. Wang, S.-S. Wong, W. Liang, M. Zanin et al., “Modified SEIR and AI prediction of the epidemics trend of COVID-19 in China under public health interventions,” Journal of Thoracic Disease, vol. 12, no. 3, pp. 165–174, Mar 2020. [Online]. Available: https://pubmed.ncbi.nlm.nih.gov/32274081

[14] S. Sanche, Y. T. Lin, C. Xu, E. Romero-Severson, N. Hengartner, and R. Ke, “High contagiousness and rapid spread of severe acute respiratory syndrome coronavirus 2,” Emerging Infectious Disease Journal, vol. 26, no. 7, pp. 1470–1477, 2020. [Online]. Available: https://doi.org/10.3201/eid2607.200282

[15] H. De-Leon and F. Pederiva, “Particle modeling of the spreading of coronavirus disease (COVID-19),” Physics of Fluids, vol. 32, no. 8, p. 087113, 2020. [Online]. Available: https://doi.org/10.1063/5.0020565

[16] N. Hoertel, M. Blachier, C. Blanco, M. Olfson, M. Massetti et al., “A stochastic agent-based model of the SARS-CoV-2 epidemic in France,” Nature Medicine, vol. 26, no. 9, pp. 1417–1421, Sep 2020. [Online]. Available: https://doi.org/10.1038/s41591-020-1001-6

[17] P. C. L. Silva, P. V. C. Batista, H. S. Lima, M. A. Alves, F. G. Guimarães, and R. C. P. Silva, “COVID-ABS: An agent-based model of COVID-19 epidemic to simulate health and economic effects of social distancing interventions,” Chaos, Solitons, and Fractals, vol. 139, p. 110088, Oct 2020. [Online]. Available: https://doi.org/10.1016/j.chaos.2020.110088

[18] M. R. Bicher, C. Rippinger, C. Urach, D. Brunmeir, U. Siebert, and N. Popper, “Agent-based simulation for evaluation of contacttracing policies against the spread of SARS-CoV-2,” medRxiv, 2020. [Online]. Available: https://www.medrxiv.org/content/early/2020/09/17/2020.05.12.20098970

[19] R. Trevethan, “Sensitivity, specificity, and predictive values: Foundations, pliabilities, and pitfalls in research and practice,” Frontiers in Public Health, vol. 5, p. 307, 2017. [Online]. Available: https://www.frontiersin.org/article/10.3389/fpubh.2017.00307

[20] Presidenza del Consiglio dei Ministri - Dipartimento della Protezione Civile. (2020) Dati COVID-19 Italia. Last accessed on 2020-10-2: https://github.com/pcm-dpc/COVID-19.

[21] Ministero della Salute. (2020) Primi risultati dell’indagine di siero-prevalenza SARS-CoV-2. Last accessed on 2020-10-2: http://www.salute.gov.it/imgs/C17notizie49980file.pdf.

[22] D. Cereda, M. Tirani, F. Rovida, V. Demicheli, M. Ajelli, P. Poletti et al., “The early phase of the COVID-19 outbreak in Lombardy, Italy,” 2020.

[23] Our World in Data. (2020) Coronavirus COVID-19 Testing. Last accessed on 2020-10-2: https://ourworldindata.org/coronavirus-testing.

[24] The Johns Hopkins Center for Health Security. (2020) Serology-based tests for COVID-19. Last accessed on 2020-10-2: https://www.centerforhealthsecurity.org/resources/COVID-19/serology/Serology-based-tests-for-COVID-19.html.

[25] C. Bonanad, S. García-Blas, F. Tarazona-Santabalbina, J. Sanchis, V. Bertomeu-González, L. Fácila et al., “The effect of age on mortality in patients with COVID-19: A meta-analysis with 611,583 subjects,” Journal of the American Medical Directors Association, vol. 21, no. 7, pp. 915 – 918, 2020. [Online]. Available: http://www.sciencedirect.com/science/article/pii/S1525861020304412

