## Supplementary materials for "A Particle-Based COVID-19 Simulator with Contact Tracing and Testing"

Askat Kuzdeuov, *Member, IEEE*, Aknur Karabay, Daulet Baimukashev, Bauyrzhan Ibragimov, and Huseyin Atakan Varol^*^, Senior Member, IEEE

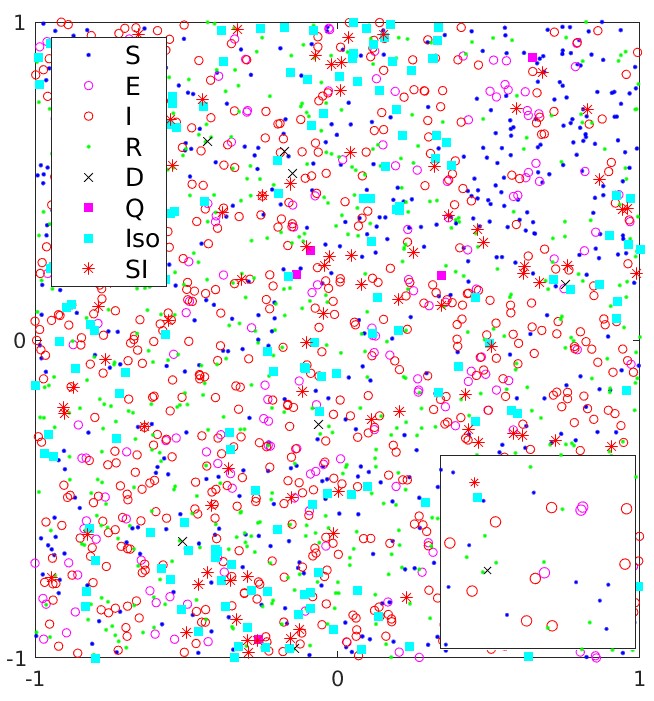

Fig. 1. Visualization of the particles on the map including their epidemic states.

I. TRANSITIONS BETWEEN EPIDEMIC STATES

In this simulator, particles move on a 2D map as shown in Fig. 1. The vector $e\in\mathbb{R}^{n}$ stores the epidemic states of all 𝑛 particles, where each element contains the epidemic status of the particle $i$, 1 ≤ $i$ ≤ $n$, and has one of these values based on the epidemic status: 0 - Susceptible, 1 - Exposed, 2 - Infected, 3 - Recovered, 4 - Dead, 5 - True Quarantined, 6 True Isolated, 7 - Severely Infected, 8 - False Isolated, and 9 - False Quarantined. Transitions between epidemic states are shown in Fig. 2. The parameters of the model are given in Table I.

At the beginning of the simulation, all $n$ particles are in the Susceptible state ($e_{i} =0$ for $i = 1,...,n$). Then, we randomly choose $n_{e}$ particles and change their epidemic status to the Exposed ($e_{\boldsymbol{r}_{k}}$= 1, for $k = 1,...,n_{e}$, where $\boldsymbol{r}\in\mathbb{R}^{n_{e}}$ is a vector of pseudorandom integers drawn from the discrete uniform distribution on the interval [1, $n$]). Then, all particles start moving on the map and come into contact with each other. A susceptible particle becomes exposed when it contacts a contagious particle (i.e., Exposed, Infected, True Quarantined, True Isolated, or

TABLE I. List of simulation parameters and their descriptions.

| **Parameters** | **Description** |
| --- | --- |
| $n_{e}$ | Initial number of exposed particles |
| $T$ | Simulation length in days |
| $x_{thr}$ | Minimum distance to transmit the disease |
| $v_{max}$ | Maximum allowed speed of particles |
| $\lambda$ | Speed gain |
| $sir$ | Daily rate of Infected/Isolated particles getting Severely Infected |
| $\gamma_{mor}$ | Severely Infected to Dead transition probability |
| $\epsilon$*_exp_* | Transmission probability of Exposed |
| $\epsilon$*_qua_* | Transmission probability of Quarantined |
| $\epsilon$*_sir_* | Transmission probability of Severely Infected |
| $t_{exp}$ | Exposure period in days |
| $t_{inf}$ | Infection period in days |
| $\beta$ | Ratio of the population using a contact tracing app |
| $\theta$ | Number of daily tests per thousand people |
| $sn$ | Sensitivity of tests |
| $sp$ | Specificity of tests |

Severely Infected). A contact between two particles is assumed to happen when the distance between them is less than $x_{thr}$ . In order to find the contacted susceptible particles, first, we need to identify contagious particles in the epidemic state vector $e$. Thus, we define $(\boldsymbol{\kappa}_{\boldsymbol{C}}\in\mathbb{R}^{n})$ to identify the indices of contagious particles, where $\kappa_{C_{i}} =1$ if a particle is contagious, otherwise $\kappa_{C_{i}} =0$. Similarly, we define vectors to identify indices of the Susceptible $(\boldsymbol{\kappa}_{\boldsymbol{S}}\in\mathbb{R}^{n})$, Exposed $(\boldsymbol{\kappa}_{\boldsymbol{E}}\in\mathbb{R}^{n})$Infected $(\boldsymbol{\kappa}_{\boldsymbol{I}}\in\mathbb{R}^{n})$,True Quarantined $(\boldsymbol{\kappa}_{\boldsymbol{TQ}}\in\mathbb{R}^{n})$,True Isolated $(\boldsymbol{\kappa}_{\boldsymbol{TI}}\in\mathbb{R}^{n})$,and Severely Infected particles $(\boldsymbol{\kappa}_{\boldsymbol{SI}}\in\mathbb{R}^{n})$ . The $\boldsymbol{\kappa}_{\boldsymbol{C}}$ is calculated based on the disease transmission probabilities as

$$\kappa_{\boldsymbol{C}} = \kappa_{\boldsymbol{I}} |(\kappa_{\boldsymbol{E}} ʘ (\boldsymbol{r} < \epsilon_{exp}))|(\kappa_{\boldsymbol{TQ}} ʘ (\boldsymbol{r} < \epsilon_{qua}))|$$

$(\kappa_{\boldsymbol{TI}} ʘ (\boldsymbol{r} < \epsilon_{qua}))|\left( \kappa_{\boldsymbol{SI}} ʘ \left( \boldsymbol{r} < \epsilon_{sev} \right) \right)$	(1)

where $\boldsymbol{r}\in\mathbb{R}^{n}$ is a vector of uniformly distributed random numbers in the range [0, 1], $ʘ$ is a pointwise multiplication operator, and | is a logical OR operator.

Then, we identify the current positions of contagious particles on the map as

${P_{C}}_{j} =P_{i}$ if $\kappa_{C_{i}} =1$ for $i = 1,...,n, j=1,\ldots m.$	(2)

where $m$ is a number of non-zero elements in $\boldsymbol{\kappa}_{\boldsymbol{C}}$, $\boldsymbol{P}\in\mathbb{R}^{nx2}$ is a matrix that stores the current positions of all particles. Then, we employ the rangesearch $(P,P_{C},x_{thr})$ function [1] of MATLAB to find particles $\boldsymbol{\kappa}_{\boldsymbol{A}}\in\mathbb{R}^{n}$ that were in contact with the contagious particles, where $\boldsymbol{\kappa}_{\boldsymbol{A}_{\boldsymbol{i}}}\boldsymbol{=}1$ if a

particle was in contact, otherwise $\boldsymbol{\kappa}_{\boldsymbol{A}_{\boldsymbol{i}}}\boldsymbol{=}0$.
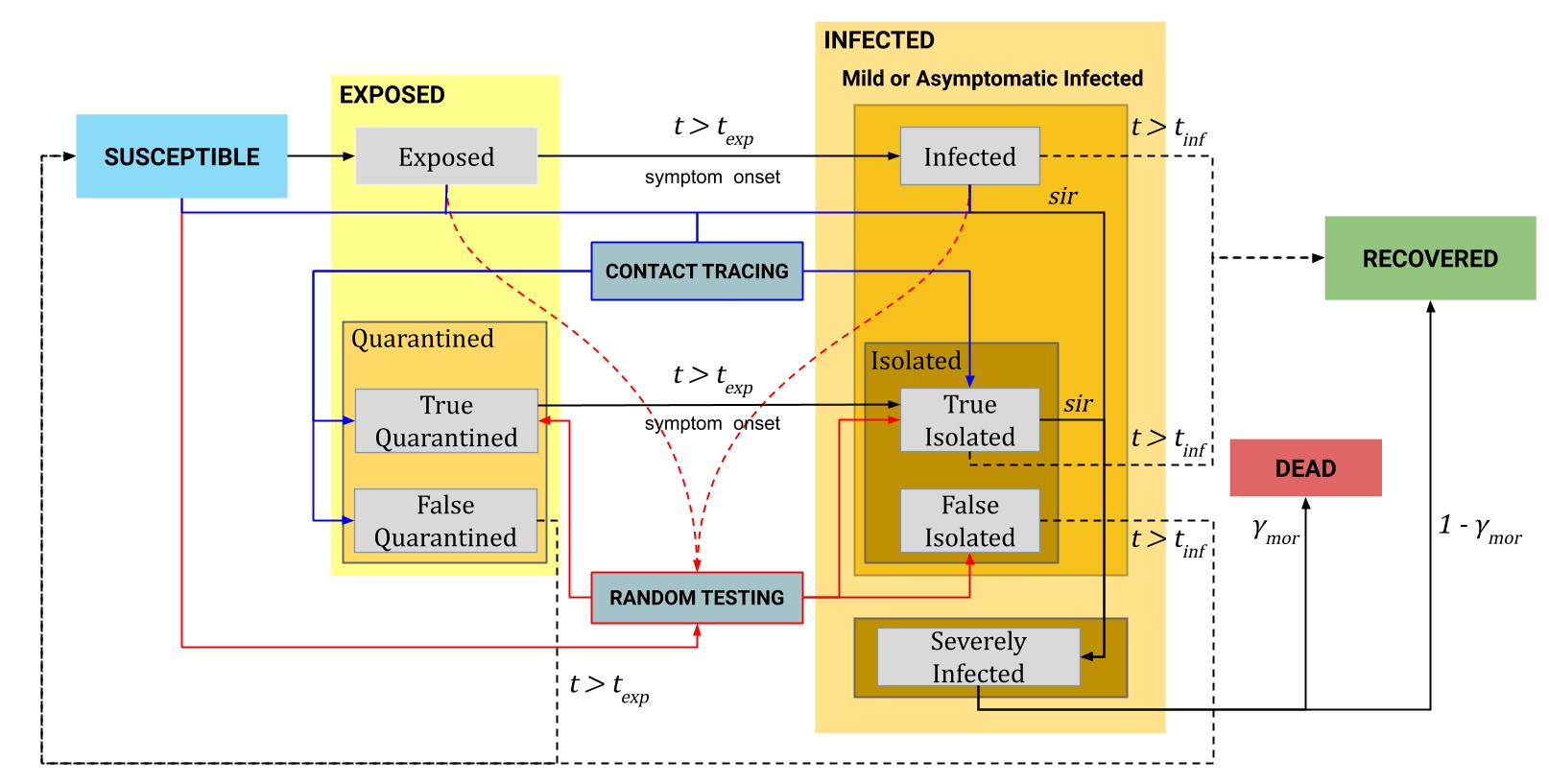
This way, we can extract the indices of the contacted susceptible particles from $\boldsymbol{\kappa}_{\boldsymbol{A}}$ as

Fig. 2. The statechart of the particle-based SEIR epidemic simulator.

$\boldsymbol{\kappa}_{\boldsymbol{S}}^{\boldsymbol{c}}\boldsymbol{=}\boldsymbol{\kappa}_{\boldsymbol{A}}\boldsymbol{\&}\boldsymbol{\kappa}_{\boldsymbol{S}}$	(3)

to change their epidemic state to Exposed and reset their epidemic time in the susceptible state:

$e_{i}=\left\{ \begin{matrix} 1, if \boldsymbol{\kappa}_{Si}^{c}=1 \\ e_{i}, otherwise. \end{matrix} \right.$	(4)

$t_{i}=\left\{ \begin{matrix} 0, if \boldsymbol{\kappa}_{Si}^{c}=1 \\ t_{i}, otherwise. \end{matrix} \right.$

The exposed particles whose epidemic time in this state exceeds the exposure period $t_{exp}$ move to the Infected state, and their epidemic time is set to zero as

$e_{i}=\left\{ \begin{matrix} 2, if e_{i}=1 \& t_{i}\geq t_{exp} \\ e_{i}, otherwise. \end{matrix} \right.$	(5)

$t_{i}=\left\{ \begin{matrix} 0, if e_{i}=1 \& t_{i}\geq t_{exp} \\ t_{i}, otherwise. \end{matrix} \right.$

Some portion of the infected particles are selected randomly and moved to the Severely Infected state during their infection period $t_{inf}$ , and regarded as positively-tested (i.e., hospitalized)

$e_{i}=\left\{ \begin{matrix} 7, if e_{i}=2 \& r_{i}\leq sir\Delta t \\ e_{i}, otherwise. \end{matrix} \right.$	(6)

${ts}_{i}=\left\{ \begin{matrix} d, if e_{i}=2 \& r_{i}\leq sir\Delta t \\ {ts}_{i}, otherwise. \end{matrix} \right.$

where $\boldsymbol{r}\in\mathbb{R}^{n}$ is a vector of uniformly distributed random numbers in the range [0, 1]; $\boldsymbol{ts}\in\mathbb{R}^{n}$ stores for each particle the date, $d$, when its COVID-19 test result was positive.

Then, the infected particles move to the Recovered state after $t_{inf}$

days:

$e_{i}=\left\{ \begin{matrix} 3, if e_{i}=2 \& t_{i}\geq t_{inf} \\ e_{i}, otherwise. \end{matrix} \right.$	(7)

Severely infected particles die with $\gamma_{mor}$ probability after $t_{inf}$ days while the rest move to the Recovered state:

$e_{i}=\left\{ \begin{matrix} 4, if e_{i}=7 \& t_{i}\geq t_{inf} \& {r_{i}\leq\gamma}_{mor} \\ 3, if e_{i}=7 \& t_{i}\geq t_{inf} \& {r_{i}>\gamma}_{mor} \\ {ts}_{i}, otherwise. \end{matrix} \right.$	(8)

The testing module randomly tests particles in the Susceptible, Exposed, and Infected states. The results of the tests might be true positive or false positive due to the test sensitivity $sn$ and specificity $sp$:

$e_{i}=\left\{ \begin{matrix} d, if (e_{i}=1 | e_{i}=2) \& {ts}_{i}=0 \& r_{i}\leq\theta sn\Delta t \\ d, if e_{i}=0 \& {ts}_{i}=0 \& r_{i}\leq\theta(1-sp)\Delta t \\ {ts}_{i}, otherwise. \end{matrix} \right.$	(9)

Particles that were correctly detected by the test are moved to the True Quarantined and True Isolated states as

$e_{i}=\left\{ \begin{matrix} 5, if e_{i}=1 \& {ts}_{i}=d \& r_{i}\leq\theta sn\Delta t \\ 6, if e_{i}=2 \& {ts}_{i}=d \& r_{i}\leq\theta sn\Delta t \\ e_{i}, otherwise. \end{matrix} \right.$	(10)

Susceptible particles that were incorrectly tested positive move to the False Isolated state:

$e_{i}=\left\{ \begin{matrix} 8, if e_{i}=0 \& {ts}_{i}=d \& r_{i}\leq\theta(1-sp)\Delta t \\ e_{i}, otherwise. \end{matrix} \right.$	(11)

Then, the contact tracing module checks the epidemic status of the particles that were in contact with the positive tested particles in the last 14 days. If the contacted particles are exposed then they transition to the True Quarantined state, if they are infected then they move to the True Isolated state. Similarly, the contacts of the Susceptible particles, which were incorrectly detected by the test, are sent to the False Quarantined state.

Particles in the True Quarantined state transition to the True Isolated when their epidemic time exceeds the exposure period $t_{exp}$:

$e_{i}=\left\{ \begin{matrix} 6, if e_{i}=5 \& t_{i}\geq t_{exp} \\ e_{i}, otherwise. \end{matrix} \right.$	(12)

$t_{i}=\left\{ \begin{matrix} 0, if e_{i}=5 \& t_{i}\geq t_{exp} \\ t_{i}, otherwise. \end{matrix} \right.$

On the other hand, particles in the False Quarantined state return to the Susceptible state after the exposure period $t_{exp}$:

$e_{i}=\left\{ \begin{matrix} 0, if e_{i}=9 \& t_{i}\geq t_{exp} \\ e_{i}, otherwise. \end{matrix} \right.$	(13)

$t_{i}=\left\{ \begin{matrix} 0, if e_{i}=9 \& t_{i}\geq t_{exp} \\ t_{i}, otherwise. \end{matrix} \right.$

Some portion of the particles in the True Isolated state are selected randomly and moved to the Severely Infected state during their infection period $t_{exp}$ :

$e_{i}=\left\{ \begin{matrix} 7, if e_{i}=6 \& r_{i}\leq sir\Delta t \\ e_{i}, otherwise. \end{matrix} \right.$	(14)

The other particles in the True Isolated state recover after $t_{inf}$ days:

$e_{i}=\left\{ \begin{matrix} 3, if e_{i}=6 \& t_{i}\geq t_{inf} \\ e_{i}, otherwise. \end{matrix} \right.$	(15)

On the other hand, particles in the False Isolated state go back to the Susceptible state when the infection period $t_{inf}$ ends:

$e_{i}=\left\{ \begin{matrix} 0, if e_{i}=8 \& t_{i}\geq t_{inf} \\ e_{i}, otherwise. \end{matrix} \right.$	(16)

$t_{i}=\left\{ \begin{matrix} 0, if e_{i}=8 \& t_{i}\geq t_{inf} \\ t_{i}, otherwise. \end{matrix} \right.$

### II. IMPACT OF THE PARTICLE VELOCITY ON THE SPREAD OF THE EPIDEMIC

The velocity of the particles plays an important role in the spread of the epidemic among the population. Higher values of $v_{max}$ lead to a faster spread of the disease. In order to show this, we conducted simulations with different values of $v_{max}=\{0.5, 0.75, 1.0, 1.25, 1.5\}$. The total number of particles $n$ is 10,000, and the initial number of exposed particles $n_{e}$ is 10. The other parameters of the simulation are shown in Table II.

For each considered maximum velocity $v_{max}$, we ran 10 simulations and took averaged results. The averaged results for all cases are summarized in Fig. 3. According to the results, the epidemic reaches the peak faster with the increased speed of particles.

TABLE II. Parameters of the simulations

| $n$ | $n_{e}$ | $x_{thr}$ | $\epsilon$*_exp_* | $\epsilon$*_qua_* | $\epsilon$*_sev_* | $t_{exp}$ |
| --- | --- | --- | --- | --- | --- | --- |
| 10,000 | 10 | 0.0001 | 0.7 | 0.3 | 0.3 | 5 |
| $t_{inf}$ | $sir$ | $\gamma_{mor}$ | $\beta$ | $\theta$ | $sn$ | $sp$ |
| 14 | 0.02 | 0.15 | 0 | 0 | 0 | 0 |

TABLE III. Simulation parameters for the canton of Geneva

| $v_{max} (Days)$ | $\lambda(Days)$ | $x_{thr}(Days)$ | $\epsilon$*_exp_* | $\epsilon$*_qua_* | $\epsilon$*_sev_* | $t_{exp}$ |
| --- | --- | --- | --- | --- | --- | --- |
| 0.02 (0-50) | 0.002 (0-50) |  |  |  |  |  |
| 0.009 [50-70) | 0.0009 [50-70) | 7.5e-5  (0-155) |  |  |  |  |
| 0.003 [70-111) | 0.0003 [70-111) | 6e-5  [155-200) | 0.7 | 0.3 | 0.3 | 5 |
| 0.004 [111-127) | 0.0004 [111-127) |  |  |  |  |  |
| 0.06 [127-155) | 0.006 [127-155) |  |  |  |  |  |
| 0.02 [155-200) | 0.02 [155-200) |  |  |  |  |  |
| $t_{inf}$ | $sir$ | $\gamma_{mor}$ | $\beta$ | $\theta$ | $sn$ | $sp$ |
| 14 | 0.01 | 0.08 | 0 | 0.5 | 0.95 | 0.99 |

In addition, the number of exposed and infected particles at the peak also grows with increased velocity. As a result, the numbers of severely infected, recovered, and dead particles increase faster. The results show that a higher velocity results in a higher number of contacts per unit time. Therefore, we changed the value of $v_{max}$, according to the epidemic timeline, to imitate different levels of the quarantine in Lecco.

III. PARTICLE-BASED SEIR SIMULATION OF THE CANTON OF GENEVA

To validate our particle-based simulator and model for the province of Lecco, we have performed simulations for the canton of Geneva in Switzerland. The official statistics [2] for this country have been shared by its government since February 25, 2020 and a number of seroprevalence studies have been performed.

The canton of Geneva is one of the most heavily hit regions of Switzerland by the COVID-19 epidemic, with 42,159 officially registered cases by January 12, 2021. However, according to the seroprevalence study [3] in the canton of Geneva from April 6 to June 30, 2020, around 7.8% of the population (38,595) had antibodies, which is seven times greater than the officially confirmed cases at that date (5,412). In order to adjust the parameters of the model for Geneva, we relied on the total cases that were corrected using the seroprevalence survey results and officially confirmed number of deaths. The total number of particles was set to 499,480 (the population of the canton) [4].

The initial number of exposed particles was set to 10. We started the simulation on January 5, 2020, in accordance with the official statistics [2]. To initialize the number of tests per thousand people, we used the averaged value 0.5 from the official statistics [2]. We adjusted parameters

such as $sir$ and $\gamma_{mor}$ while the other parameters were the same as for the province of Lecco. All parameters of the simulation are provided in Table III. To imitate the lockdown in the canton of Geneva, we decreased the maximum speed of particles $v_{max}$ and $\lambda$, in accordance with the epidemic timeline. The detailed timeline of policies in the canton of Geneva is provided in Table IV.

We ran 10 simulations, and the averaged results are shown in Fig. 4. The results show that by tuning only a few parameters of the calibrated model we can fit data for other countries. The model also predicted the peak of the epidemic accurately, because, according to the official data, the number of cases increased starting from the middle of March and remained high until the middle of April. In addition, the model predicted the second wave of the epidemic after 180 days. The same can be observed in the official data (see Fig. 5).

| **Day** | **Date** | **Event** |
| --- | --- | --- |
| 0 | 5/1/2020 | Start of the simulation. |
| 50 | 25/2/2020 | The COVID-19 data repository was launched [3]. First confirmed case. |
| 70 | 16/3/2020 | Lockdown in the canton. |
| 111 | 26/4/2020 | 1st stage of lockdown easing.  Hospitals for all purposes are open. |
| 127 | 11/5/2020 | 2nd stage of lockdown easing Shops, markets are open. |
| 155 | 8/6/2020 | 3rd stage of lockdown easing Schools are open. |
| 200 | 19/7/2020 | End of the simulation. |

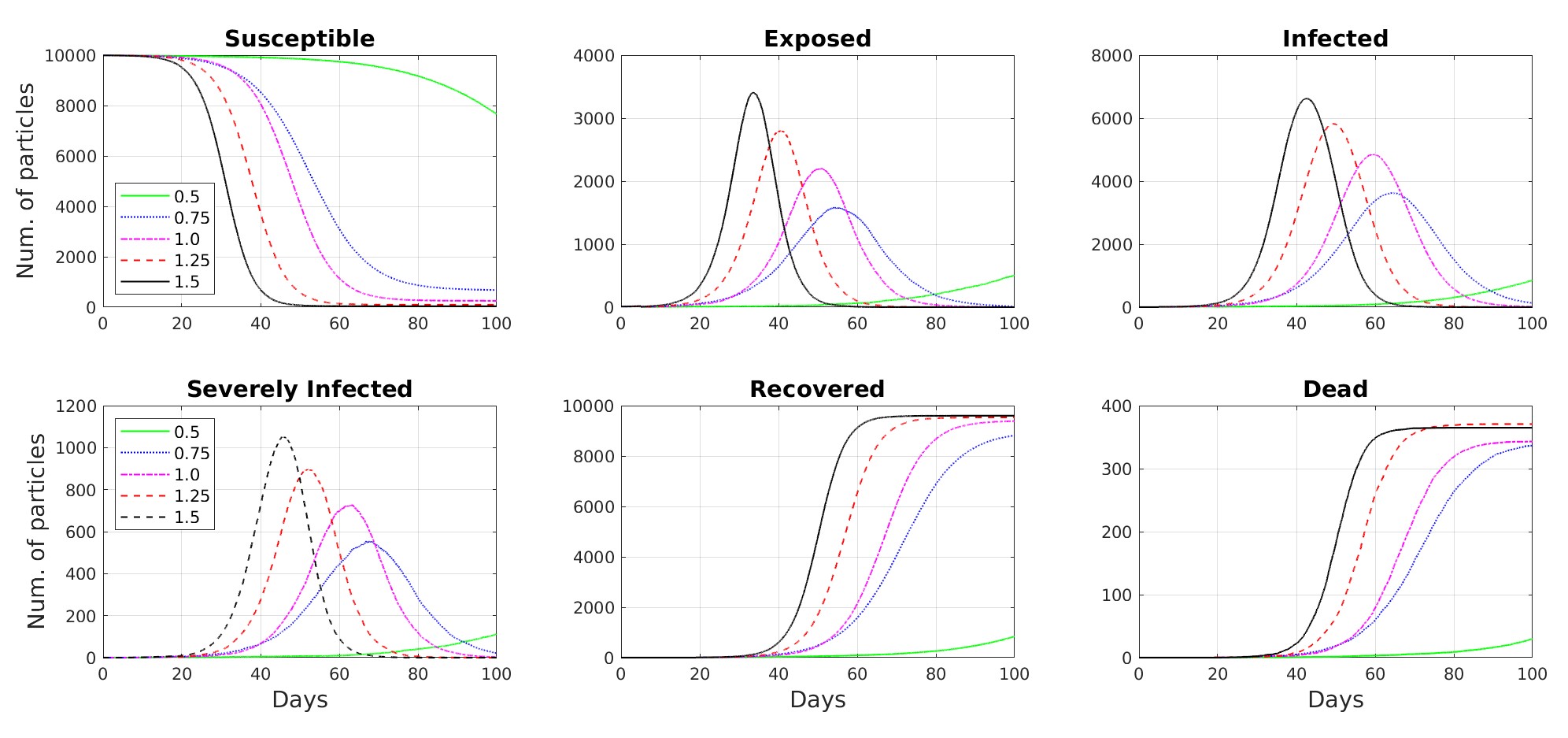

TABLE IV. COVID-19 policies timeline in the canton of Geneva [5]

REFERENCES

[1]	MATLAB.	rangesearch.	Last	accessed	on	2021-02-2: https://www.mathworks.com/help/stats/rangesearch.html#mw 51f765d0-dc9e-42f3-83dd-472717aa3f57.

[2]	Statistisches Amt. (2021) Data COVID-19 Swiss Cantons and Principality of Liechtenstein. Last accessed on 2021-01-15: https://github.com/ openZH/covid 19.

Fig. 3. Simulation results for different values of $v_{max}$

[3]	A. Richard, A. Wisniak, J. Perez-Saez, H. Garrison-Desany, D. Petrovic, G. Piumatti et al., “Seroprevalence of anti-SARS-CoV-2 IgG antibodies, risk factors for infection and associated symptoms in Geneva, Switzerland: a population-based study,” medRxiv, 2020. [Online]. Available:

https://www.medrxiv.org/content/early/2020/12/18/2020.12.16.20248180

[4]	Data Commons. Canton of Geneva. Last accessed on 2021-01-15: https: //datacommons.org/place/nuts/CH013.

[5]	CNBC. Switzerland to start easing Covid-19 restrictions from April

27. Last accessed on 2021-01-10: https://www.cnbc.com/2020/04/16/ switzerland-to-start-easing-covid-19-restrictions-from-april-27.html.

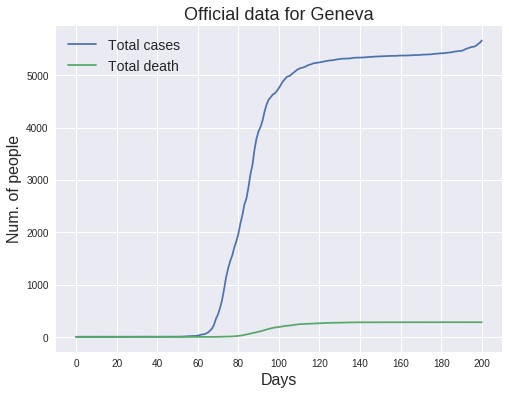

Fig. 4. The averaged results of ten simulations for the canton of Geneva. The upper plot shows the states of the epidemic simulation versus time. The dates of important NPIs are shown by vertical dashed lines and listed in Table IV. One standard deviation around the average Total case curve is shaded in blue. The bottom plot compares the average number of deaths in the simulation with the actual number of deaths due to COVID-19. This plot also shows the simulation results for the number of severely infected particles as well. One standard deviation around the average Dead state curve is shaded in grey.

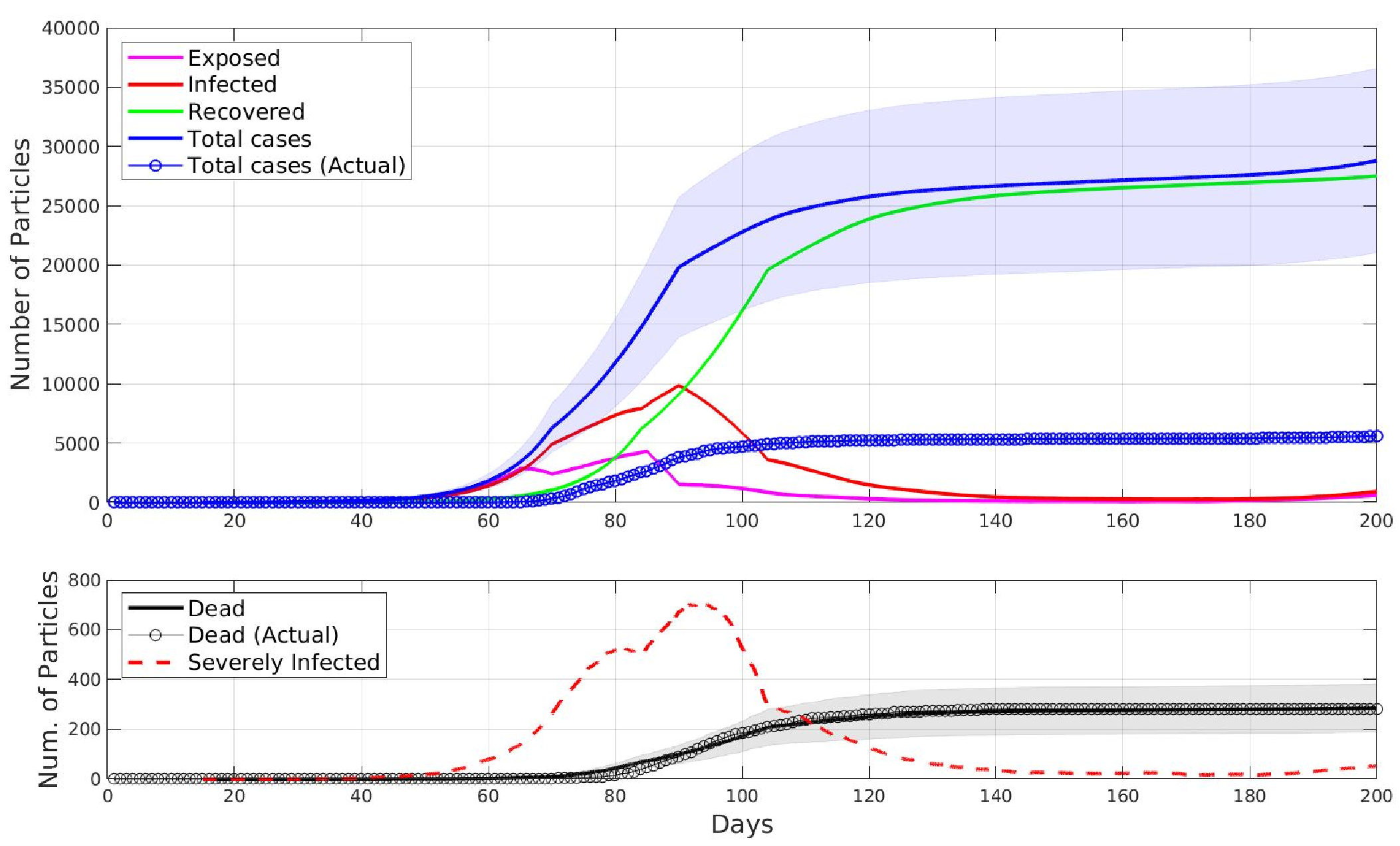

**22**

**7/2020**

**/**

**5**

**/**

**1/2020**

**25**

**/**

**2/2020**

**3/2020**

**/**

**16**

**4/2020**

**/**

**26**

**/**

**5/2020**

**11**

**8**

**6/2020**

**/**

Fig. 5. Actual COVID-19 data for the canton of Geneva from 5 January 2020 to 11 August 2020.
